# A New Dynamic Echodefecography (EDF) Established by Using BK Ultrasound 8838 Transducer and its Testing Results Comparison with X-ray Defecography

**DOI:** 10.1101/2020.04.06.20049551

**Authors:** Heiying Jin, Chunxia Zhang

## Abstract

**Objective:** To describe a new dynamic echodefecography(EDF) using BK ultrasound 8838 transducer and compare its testing results with X-ray defecography.

**Method:** The BK 8838 ultrasound probe is used to evaluate the static 3D scan, dynamic 3D scan and dynamic 2D scan of pelvic floor and compare its testing results with X-ray defecography and defined its value to evaluate the pelvic floor disease.

**Results:** Fifty-seven patients were studied (24 male and 33 female). Forty seven patients were diagnosed as anismus by EDF and 46 patients were diagnosed as anismus by X-ray defecography. Sixteen patients were diagnosed as rectocele by X-ray defecography, among which eight were classified as mild(6-15mm), 4 as moderate(16-30mm) and 4 as severe(over 30mm).Fourteen patients with constipation and 2 patients with anal pain were diagnosed as intussusception by EDF, but only 3 patients were diagnosed as intussusception by X-ray defecography. Two patients with constipation were diagnosed as perineal descent by EDF and none by X-ray defecography. Two patients were diagnosed as enterocele by EDF as well as X-ray defecography.

**Conclusion:** The EDF established by BK 8838 ultrasound probe can show clear anatomy and real time movement of pelvic floor muscle. The EDF is more sensitive to the diagnosis of intussusception, perineal descent(PD) and anal spincter defect than X-ray defecograpgy. For anismus,rectocele and enterocele,the diagnosis results are comparable between EDF and X ray defecography. Further study is needed to determine its clinical values to evaluate the pelvic disease.

## Introduction

Three-dimensional(3D) anorectal ultrasound is an important method for the diagnosis of pelvic floor dysfunction. Mura-dRegadas SM et al.^[1-3]^ reported dynamic echodefecography(EDF) by 2050 or 2052 anorectal transducer (B-K Medical, Herlev, Denmark) to evaluate the pelvic floor disease inctal 2008 (Surg Endosc. 2008), which plays an important role in the diagnosis of pelvic floor dysfunction and outlet obstructive constipation, and has many advantages such as being minimally invasive, inexpensive, well tolerated and X-ray exposure avoided. The EDF can show clear floor muscle and anatomic structure and is considered as ideal method to diagnose the pelvic floor disease compared with X-ray defeography ^[4-6]^.

Recently, based on the principle of EDF established by Murad-Regadas SM^[5-6]^, we used the BK 8838 transducer (Endocavity 3D 8838)to carry out dynamic EDF detection, which was easier to operate and can get more clear images and real time movement of pelvic floor muscle. The aims of the study are to describe this new dynamic echodefecography(EDF) estabilished by using the BK ultrasound 8838 transducer and compare the results with X-ray defecography.

### Patients

From Feb. 2019 To Aug 2019, 57 patients in Department of colorectal surgery, the second affiliated hospital of Nanjing University of TCM, were performed dynamic EDF with BK 8838 transducer as well as X-ray defecography. Twenty four patients were male and 33 were female. The medium age was 62(ranged from 30 to 77yrs). Forty-four patients were clinical diagnosed as obstructive constipation, eight patients were anal pain and 5 patients were fecal incontinence. All the patients signed the informed consent form. All the patients performed the dynamic EDF as well as X-ray defecation. X-ray defecation procedure and diagnosis criteria based on the literature reports^[7]^.This study has been approved by the hospital ethics committee of The second affiliated Hospital of Nanjing University of Chinese Medicine(201902001). All patients have informed consent.

## Method

### The Imaging principle of dynamic EDF byBK 8838 probe

The 8838 probe has an ultrasonic transducer which can rotate 360 degrees counterclockwise and can perform 2D and 3D scanning. When it scans in 2D, a sagittal plane (linear array in posterior or anterior) or a coronal plane image (linear array in left or right side) can be obtained. If the patient is asked to perform the action of strain or defecation, the real-time pelvic floor muscle movement can be observed clearly. The 8838 transducer also has a 3D mover. During the 3D mode scanning, the linear array will move counterclockwise and can get lots frames sagittal or coronal cross-sectional image and then the 3D image can be obtained by 3D software imaging. It takes about 48 seconds to complete the whole 3D scanning process (Figure1A,B,C).

**Figure1:**
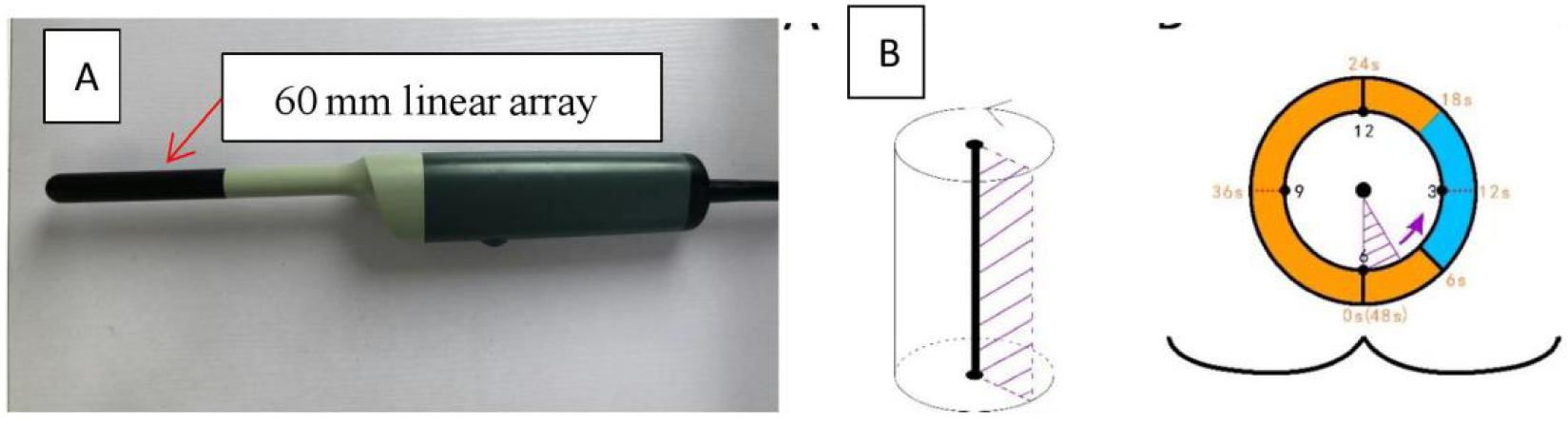
BK 8838 transducer(A:Endocavity 3D 8838.B 8838 transducer operating diagram.)

### Procedure for dynamic EDF

1. Preoperative preparation: enema with sodium phosphate enema;
2. Patient’s position: Semi-reclining lithotomy position(Figure2);
3. Insert 50-100 ml and 10-20 ml of ultrasound gel into the rectal ampulla and the vagina respectively, to achieve contrast and get better images. The scanning frequency was 9-12 Hz. The 8838 transducer is put into the anus and rectum. The upper margin of reached prostate or the cervix, which was 6 cm from anal verge. (Figure3, Figure4A, B)

**Figure2:**
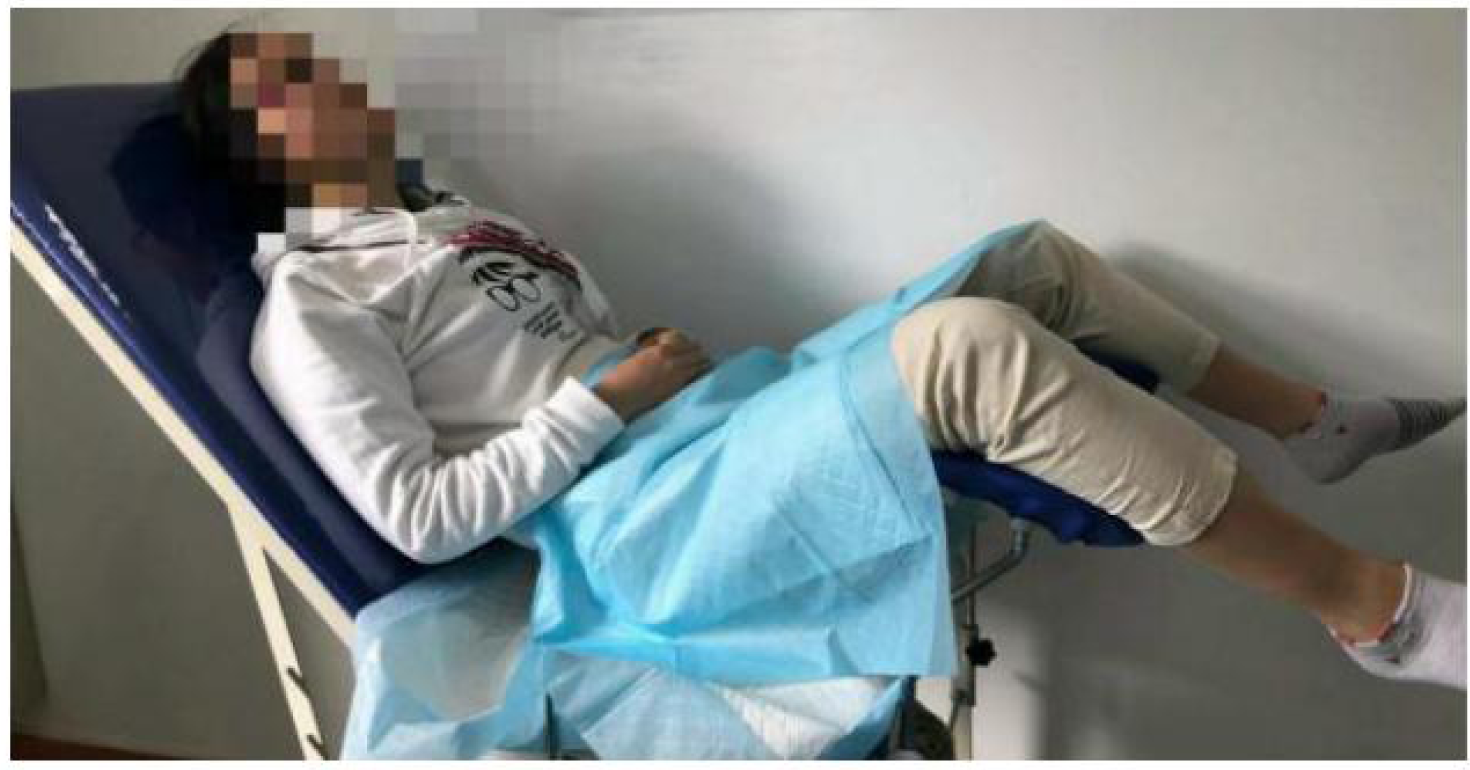
Semireclining Lithotomy(SRL)

**Figure3:**
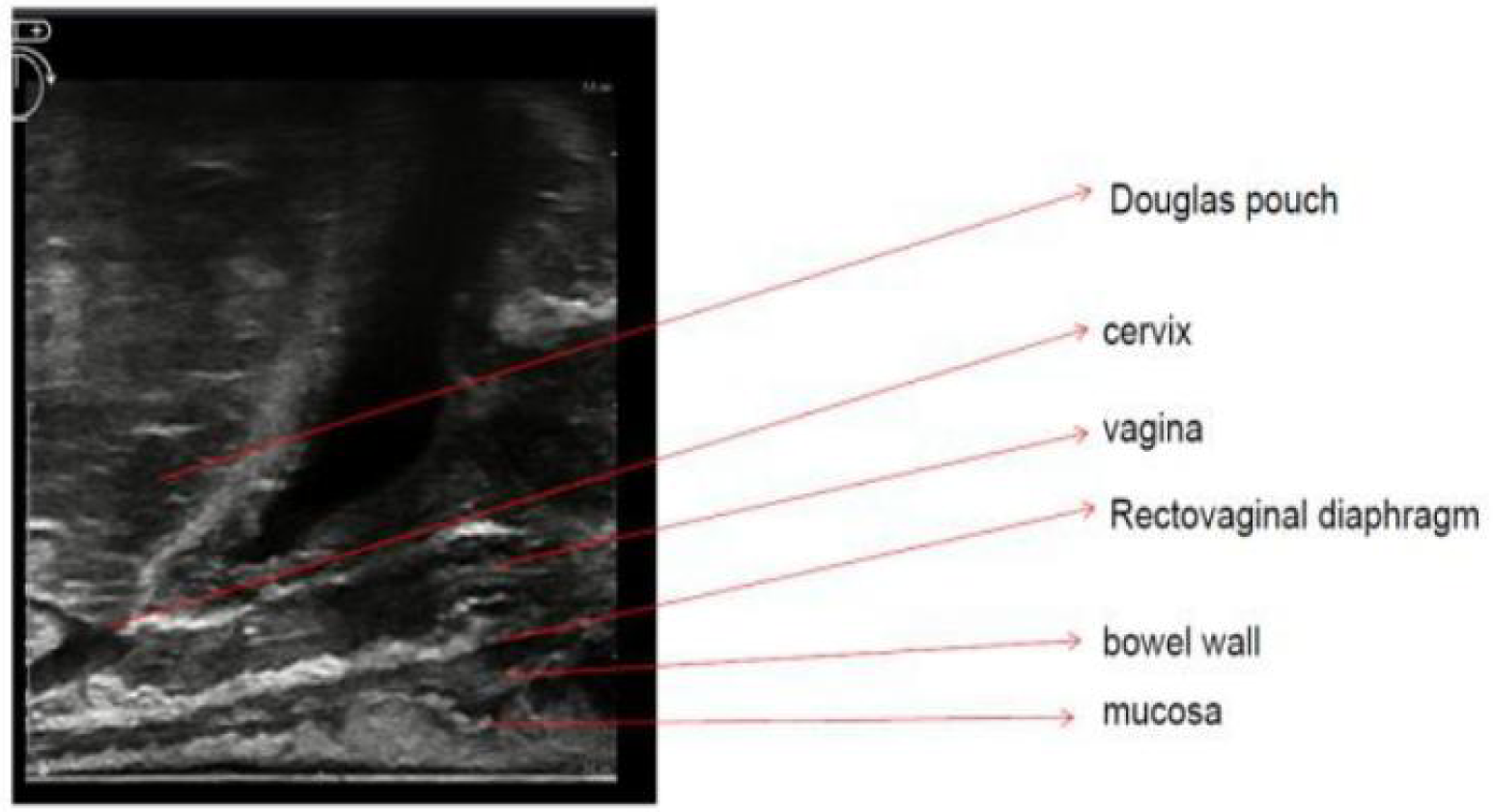
the structure of anterior wall of rectum and vagina(2D scanning)

**Figure4.**
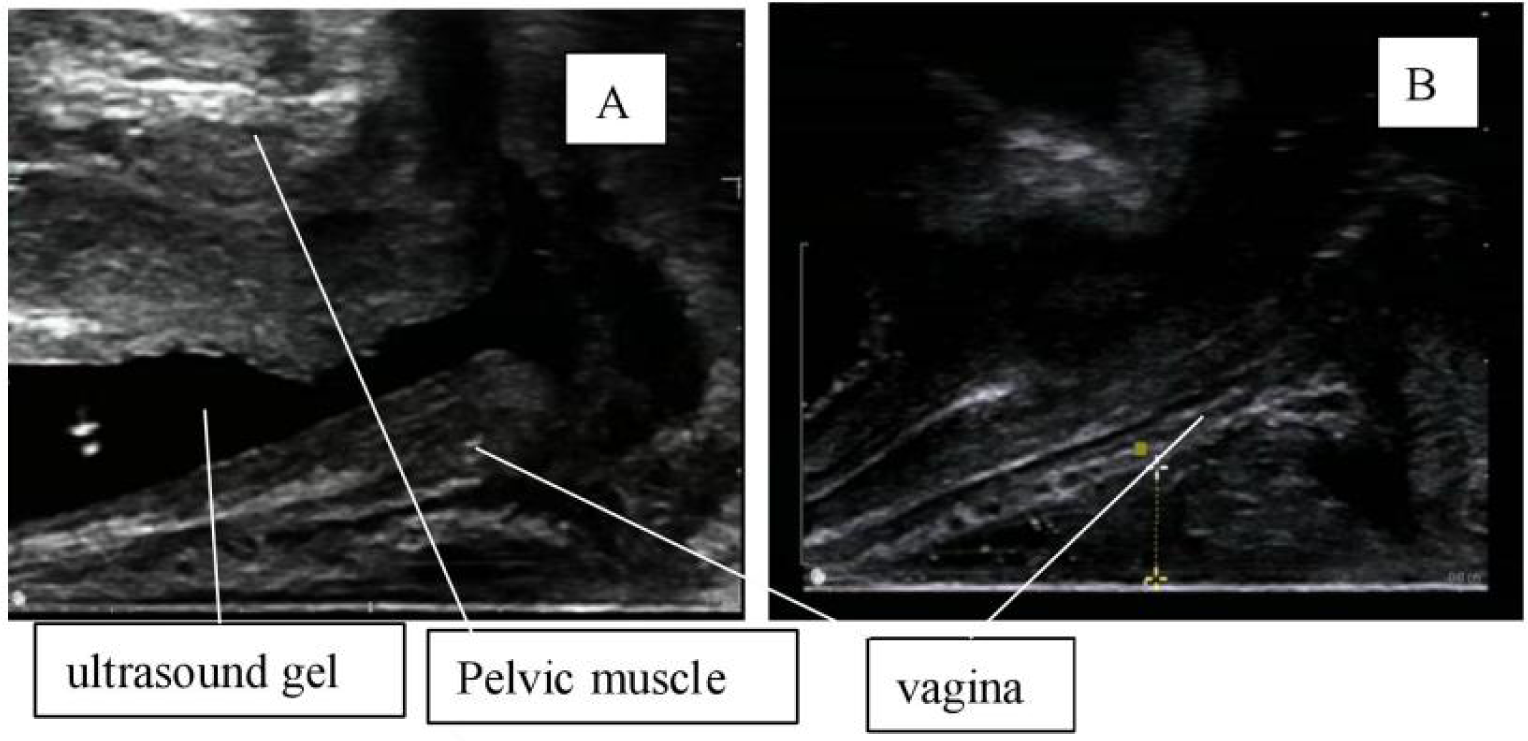
The defference of image the vagina with and wichout ultrasound gel (A:with ultrasound gel in vagina, which showed very clear pelvic floor muscle;B without gel in vagina, which showed very clear pelvic floor muscle)
4. Three-dimensional(3D) scanning is performed to observe the structure of the rectal and anal canal, the anorectal angle at rest (the angle between the puborectalis and external sphincter lines and the vertical line of the anal canal), the position of the puborectalis and anal sphincters, the pelvic floor muscles, the prostate, the uterus, and the Douglas pouch (Fig5).

**Figure5:**
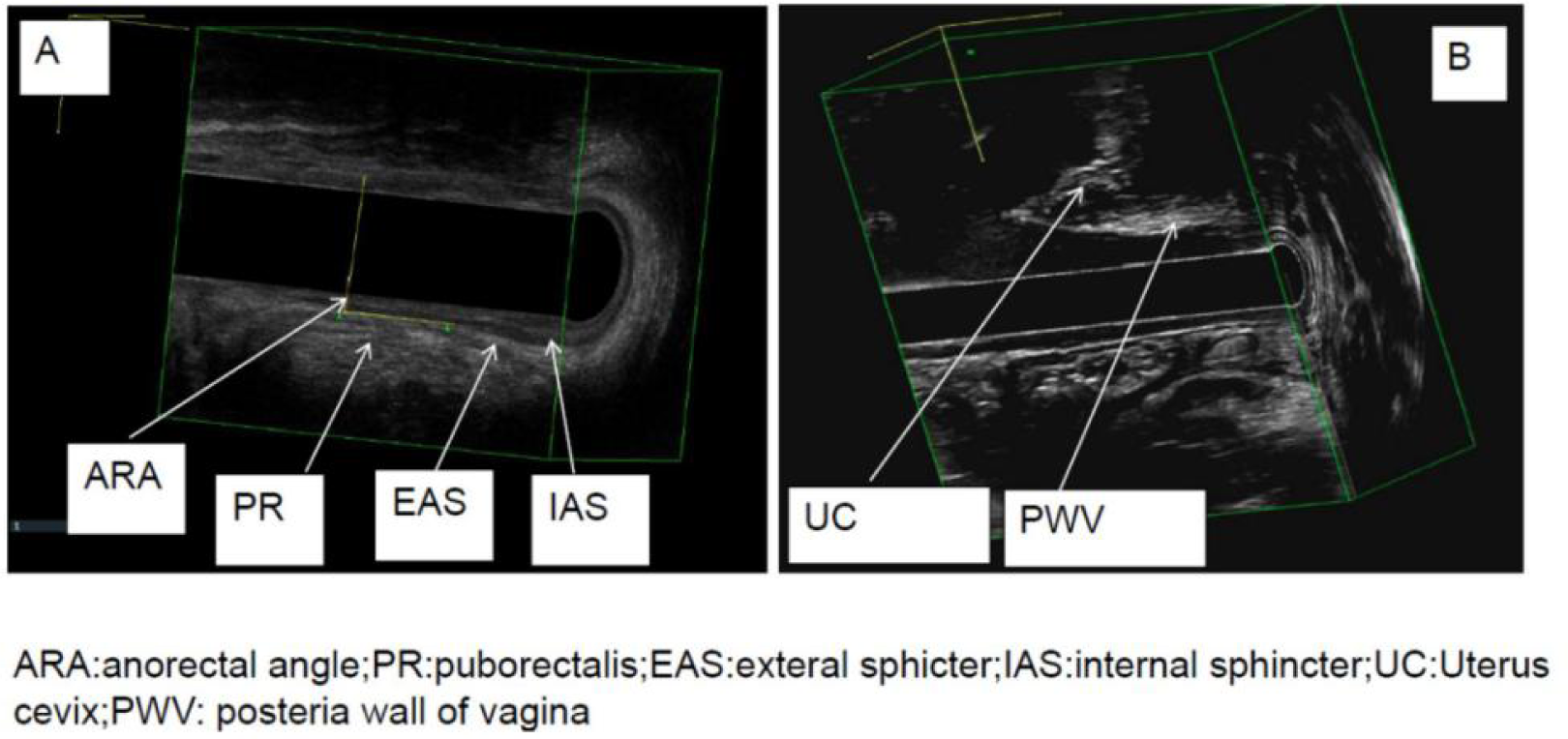
The structure of the rectum, anal and vagina(3D at rest)
5. Dynamic 3D scanning, the patients are asked to rest for 6s and then maximum defecation strain for over 12s and then rest for 30s. This scan identified and quantified the depth of rectocele as well as intussusception, sigmoidocele/enterocele, cystocele and puborectalis descent (Figure6).

**Figure6:**
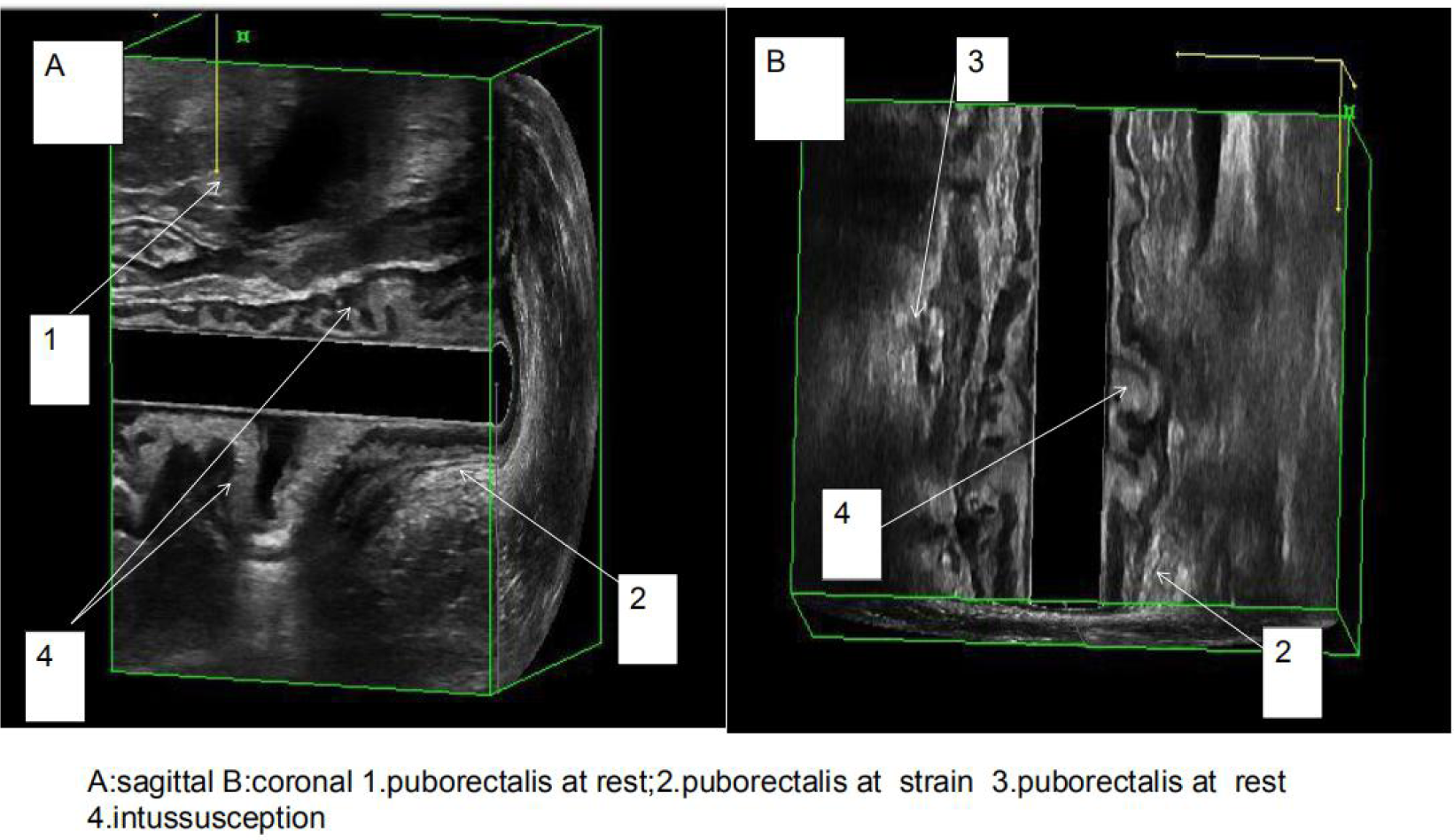
Dynamic 3D scanning
6. The 2D real-time scanning (video in supplement material):

Anterior scanning: The linear array of 8838 transducer is faced anterioly(12 o’clock direction) and the anterior wall of the intestine and anal canal is closely attached. The puborectalis are marked firstly, and then the patients are asked to defecate forcefully and the process video is recorded. In real-time video, the movement of pubrectalis, the open or closure of anorectal angle, the movement of sphincter and rectal wall, the rectocele as well as intussusception, the sigmoidocele/enterocele, the cystocele can be observed dynamically.

Left and right side scanning: The linear array of 8838 transducer is faced to the left(3 o’clock direction) and the right(9 o’clock direction). The pelvic floor muscles are marked first, then the patient is asked to defecate forcefully and the process video is recorded. The movement of bilateral pelvic floor muscles can be record. The enterocele, rectal wall and rectal mucosa at the bilateral peritoneal reflex are observed dynamically.

Posterior scanning: The linear array of 8838 transducer is faced posteriorly(6 o’clock direction). The presacral space is examined firstly and then the patient is asked to defecate forcefully and the process video was recorded. The rectal wall and mucosa are observed dynamically.

### Define the results ^[4-6]^

1. EDF Anismus: more than one degree of closure of the anorectal angle during defecation, as compared to rest (Figure7).

**Figure7:**
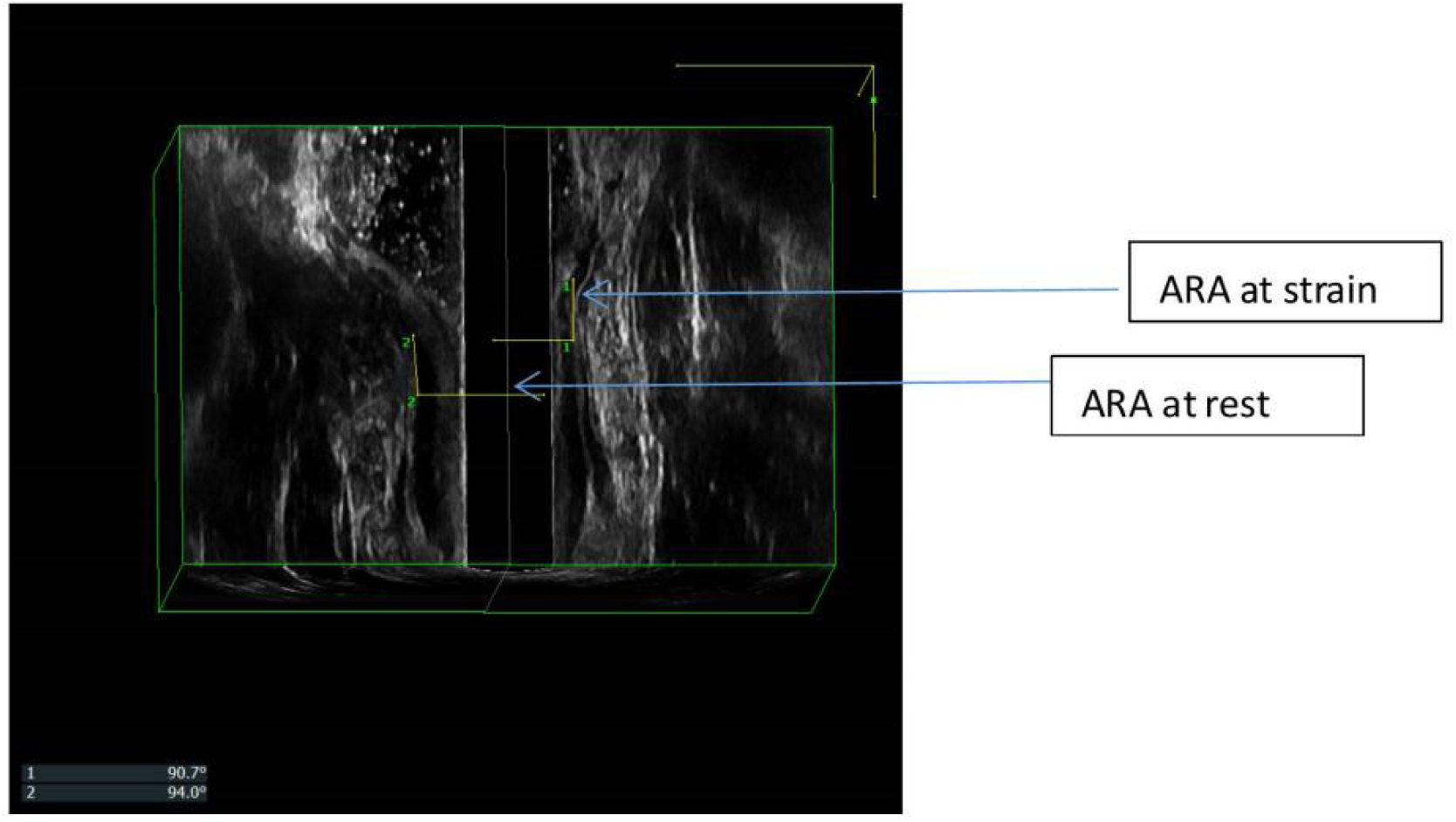
Anorectal angel(ARA) during defecation and rest
2. EDF rectocele: after injecting 50-100 cc of ultrasound gel into the rectum and during defecation straining, an anomalous hypo-echoic image is observed which was not visible at rest. This image corresponds to a hernia of the anterior wall of the rectum filled with ultrasound gel. Graded classification is defined according the X-ray defecography, which is applied as the “gold standard”.
3. EDF intussusception: The image can appreciate a “finger like shape” at the anterior or posterior level, which is composed by all the layers of the rectum. The deeper the invagination, the clearer the image.
4. EDF perineal descent (PD): puborectalis descent during maximum strain higher than 2.5 cm
5. EDF enterocele: observation of the intestinal loop between the vagina and the rectum during defecation straining.
6. Anal sphincter defect: describe the anal defect according to the site and the angle of the sphincter defect.

## Results

Dynamic EDF were performed on all 57 patients and then X-ray defecography was performed at the second and the third day.We compared the results from dynamic EDF with those from X-ray defecographyfor diagnosis of pelvic floor disease and adjusted the diagnosis criteria according to the X-ray defecography outcome.

Anismus: 47 patients were diagnosed as anismus by EDF and 46 patients were diagnosed as anismus by X-ray defecography. The results were comparable between EDF and X-ray defecography.

Rectocele: Sixteen patients were diagnosd as rectocele by X-ray defecography[7]. Eight were classified as mild (6-15mm), 4 as moderate(16-30mm) and 4 as severe (over 30 mm)[8-9]. According to this criteria, the standard for mild by EDF was 4.0-7.9mm, for moderate was 7.9-14.2mm and for severe was over 14.2mm.(Figure8)

**Figure8:**
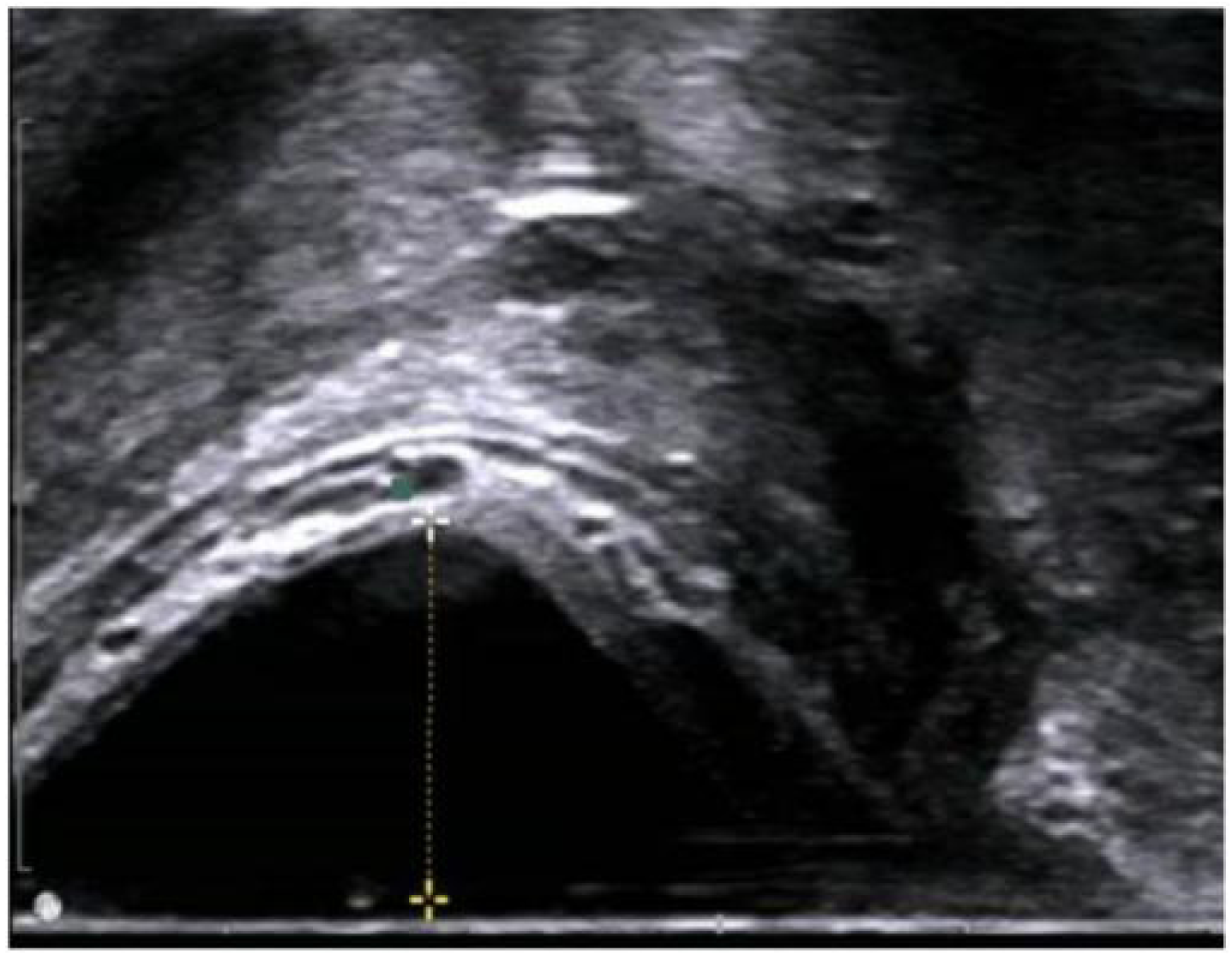
rectocele

Intussusception: Fourteen patients with constipation and 2 patients with anal pain were diagnosed as intussusception by EDF, but only 3 patients were diagnosed as intussusception by X-ray defecography.

Perineal descent (PD): Two patients with constipation were diagnosed as perineal descent by EDF and no one by X-ray defecography.

Enterocele: Two patients were diagnosed as enterocele by EDF as well as X-ray defecography.

Anal sphincter defect: Five patients with fecal incontinence were found with sphincter defect by EDF. Four were external were sphincter defect and one was internal sphincter defect.(Figure9)

**Figure9:**
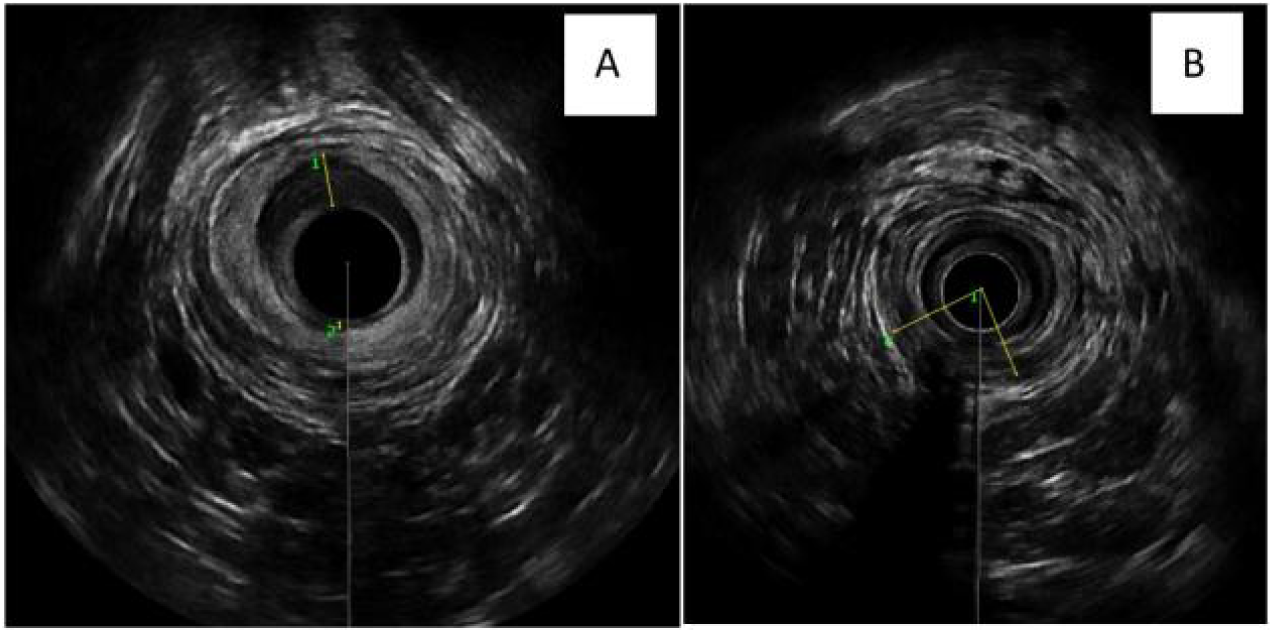
Anal spincter defect(A:internal sphincter defect;B external sphincter defect)

## Discussion

The EDF can show clear floor muscle and anatomic structure and is considered as an ideal method to diagnose the pelvic floor diseases compared with X-ray defeography.^[10-14]^. Xue et al. ^[15]^ used EDF to study functional anal pain. It was found that abnormal contraction and hypertrophy of puborectalis muscles were the main causes of functional anal pain.

However,some shortcomings exist in the BK 2050/2052 anorectal transducer when performing dynamic EDF^[14]^. The BK2050 or 2052 anorectal transducer has a rotating bicrystal at the top, which is scanned radially in a 90-degree plane at the rate of 4-6 cycles per second to form 360-degree cross-sectional scanning images. The position of the bimorph can be moved by a control button on the handle to scan different parts of the anal and rectum. The BK2050 or 2052 anorectal transducer has a 3D mover. During the 3D mode scanning, the ultrasonic bicrystal is scanned from the head to the tail end, gradually complete the different parts of the 2D scanning, and finally through data integration, form a 3D image. It takes about 60 to 70 seconds to complete the whole 3D scanning process^[1-4]^.

According to the principle of BK 2050 or 2052 anorectal transducer, we analyze its theoretical and practical defects. The dynamic EDF established by BK 2050 or 2052 anorectal transducer^[1-6]^is divided into four scans. We analyze its shortcomings step by step.

Scan 1 (at rest) is to verify the anatomical integrity of the anal sphincters. At this scan, the ideal result can be achieved by scanning in static state.

The Scan 2 includs 35s rest, maximum defecation strain over 20 s and 15 s rest again to evaluate the movement of the puborectalis muscle and the external anal sphincter during straining, identifying normal relaxation, non-relaxation or paradoxical contraction (anismus)^[2-4]^. The purpose of scan 2 is to understand the initial state of puborectalis muscle and rectal angle of anus by static scanning for 35 seconds, and then asking the patient to keep defecation strain for over 20s to push the puborectal muscle at the lowest point when the bicrystal moves to the right plane to scan the puborectal muscle, which can determine whether there is abnormal adduction of puborectal muscle by comparing the change of anorectal angle in static position and in defecation strain position. However, practically patients often can not accurately maintain for over 20s seconds, and therefore it is difficult to accurately determine whether the puborectalis muscle is at the lowest point when the bicrystal reaches the right plane, which leads to examination errors or even mistakes. In addition, the 3D images are formed by lots of frames of cross-sectional static images, so the ideal imaging should be achieved in static station and dynamic scanning often makes the image blurred, indistinct, and too inaccurate to read

In scan 3, the patients are re-asked to rest 3s followed by maximum defecation strain to quantify the perineal descent (puborectalis muscle (PR) descent) by measuring the distance between the position of the proximal border of the puborectalis muscle at rest and the point to which it is displaced by maximum straining^[2-4]^. Perineal descent B2.5 cm is classified as normal, while Perineal descent over 2.5 cm is classified as excessive. Thus, there are some problems about scan2 and scan 3, which causes the image to be blurred, indistinct and inaccurate to read

In scan 4, 150 ml and 50 ml of ultrasound gel into the rectal ampulla and the vagina respectively, to achieve the contrast. Patients are asked to rest for 35s and then maximum defecation strain for over 20s and then rest for 15s^[2-4]^. This scan identifies and quantifies the depth of rectocele as well as intussusception, sigmoidocele/enterocele (grade II or III) and cystocele. The image from scan 2 is as blurred and indistinct as the one from scan 3. The key problem is that the 3D ultrasound images are combined by two-dimensional cross sections, so the EDF established by 2052 or 2050 transducer is actually a static image combination, which cannot be considered dynamic at all and can not reflect the real-time movement of the pelvic floor. In clinical practice, X-ray defecography can not obtain real-time dynamic movement images.

.The 8838 probe has different image principles by 2052 or 2050 probe. In our study, we compared the outcome by EDF with the one by X-ray defecography. For the diagnosis of anismus, rectocele and enterocele, the outcome was comparable beteween EDF and X-ray defecography. For the diagnosis of intussusception, perineal descent(PD) and anal spincter defect, the EDF was more sensitive that X-ray.

Dynamic defecography has many advantages in helping us to understand the pelvic floor movement^[16-18]^. Some authors used dynamic MR defecography for evaluating the pelvic floor disease but MR defecography is expensive and not very convenient clinically. The dynamic EDF by 8838 probes can capture real-time video and get more information of pelvic floor muscle movement.

In addition,Murad-Regadas SM et al^[1-3]^ reported that EDF was examined in lateral decubitus position. Because the lateral decubitus position does not conform to the normal pelvic floor movement posture and defecation position. some patients do not adapt to the lateral decubitus position, which may result in abnormal contraction or relaxation of the puborectal muscles.

In conclusion, the EDF established by BK 8838 ultrasound probe can show clear anatomy and real time movement of pelvic floor muscle. The EDF is more sensitive to the diagnosis of intussusception, perineal descent(PD) and anal spincter defect than X-ray defecograpgy. For anismus,rectocele and enterocele,the diagnosis results are comparable between EDF and X ray defecography.Further study is needed to determine its clinical value to evaluate the pelvic disease.

## Data Availability

the availability of all data referred to in the manuscript and note links below.

